# Diagnostic accuracy of a high-throughput multiplex immunoassay for the detection of Mpox virus infection and MVA-BN vaccination up to two years after exposure

**DOI:** 10.1101/2025.06.12.25329495

**Authors:** Joachim Mariën, Christophe Van Dijck, Nicole Berens-Riha, Elisabeth Willems, Luca M. Zaeck, Scott Jones, Bethany Hicks, Ashley Otter, Jenifer L. Yates, Danielle T Hunt, William T. Lee, Stefanie Bracke, Jacob Verschueren, Fien Vanroye, Evi Bosman, Natalie De Cock, Bart Smekens, Leen Vandenhove, Odin Goovaerts, Anke van Hul, Patrick Soentjens, Arnaud Marchant, Marjan Van Esbroeck, Koen Vercauteren, Wim Adriaensen, Rory D. de Vries, Kevin Ariën, Laurens Liesenborghs

## Abstract

**Background:** Mpox, caused by mpox virus (MPXV), has gained global attention following the 2022 Clade

IIb outbreak and the emergence of two novel Clade I lineages in 2023 and 2024. In this context, accurate and high-throughput detection of MPXV-specific antibodies has become essential for surveillance programs, diagnosis and vaccine trials. To address this need, we validated the long-term diagnostic accuracy of an mpox multiplex IgG immunoassay (called MpoxPlex) that measures IgG responses to 11 orthopoxvirus antigens.

**Methods:** The MpoxPlex assay was validated in a Belgian cohort of mpox patients (n=226) and MVA-BN vaccine recipients (n=194), who were followed up for two years post-exposure. We used receiver operator characteristic (ROC) analysis to assess diagnostic performance at multiple time points post-infection or vaccination. To explore the assay’s broader utility, we also investigated correlations between antigen-specific IgG responses and neutralizing antibody titers. To further optimize antigen combinations, we applied a machine learning (random forest) algorithm.

**Results:** In individuals without prior smallpox vaccination, the MpoxPlex assay accurately detected mpox infection up to two years post-infection, achieving 82% sensitivity and 95% specificity when multiple antigens were combined. In contrast, diagnostic performance was low in previously smallpox or MVA-BN vaccinated individuals, with sensitivity dropping below 50% already after six months. Among MVA-BN–vaccinated individuals, IgG titers waned within one year. Most antigen-specific IgG responses correlated strongly with neutralizing antibody titers, supporting their relevance in immune monitoring.

**Conclusions:** The MpoxPlex assay exhibits high sensitivity and specificity for detecting prior mpox infection in unvaccinated individuals, with reliable performance up to two years post-infection. This makes it well-suited for both retrospective diagnosis and large-scale serosurveillance. Its strong concordance with neutralizing antibody titers further supports its potential for monitoring long-term vaccine-induced immunity. However, antibody levels following MVA-BN vaccination waned within one year, and prior vaccination status should be carefully considered when interpreting results.

## Background

Mpox, formerly known as monkeypox, is a zoonotic disease caused by mpox virus (MPXV), a member of the Orthopoxvirus genus^1^. Two clades are known to cause disease. Clade I, which originated in Central Africa, and Clade II, with origins in West Africa. Historically, these two caused outbreaks in remote regions of Central and West Africa mainly through zoonotic spillover from an unidentified animal reservoir^2^. This changed when, in 2022, subclade IIb was able to spread efficiently in worldwide sexual networks, leading to a global outbreak, which disproportionally affected gay, bisexual and other men who have sex with men(GBSMS)^3^. In 2023-24, two other lineages associated with sustained human-to-human transmission emerged in the Democratic Republic of Congo; one in Eastern DRC, named Clade Ib, the other in Kinshasa, a new lineage of Clade Ia^4,5^. The rapid spread of new lineages may be partly attributed to the halt of smallpox vaccination following its eradication in 1980, which left a growing part of the population susceptible^6,7^. In response to the 2022 outbreak, targeted campaigns using the Modified Vaccinia Ankara–Bavarian Nordic (MVA-BN) vaccine were therefore implemented in at-risk communities^8^. However, our group and others recently demonstrated a significant decline in IgG and neutralizing antibody levels already one year after MVA-BN vaccination^9–11^. Mpox therefore remains a global public health concern for which improved diagnostic and surveillance capabilities are highly needed^12–15^.

Traditional serological assays have several limitations when used for mpox diagnostics^12^. Virus neutralization tests, while still regarded the gold standard for vaccine studies, are expensive, time-consuming, and require specialized laboratory infrastructure^11,12^. ELISA assays offer a more affordable and high-throughput option. However, they assess only one analyte (whole pathogen or single antigen) at a time, making it difficult to rule out cross-reactivity with antibodies against other pathogens^6,7^. This poses a particular challenge in epidemiological studies, where assays must be highly accurate to generate population-level insights. A high accuracy is especially crucial in low-incidence settings where false-positive results can significantly bias surveillance outcomes^15^. There is thus an urgent need for serological assays that are not only sensitive and highly specific, but also scalable and suitable for use in resource-limited settings where mpox is most prevalent^14^. In this context, multiplex immunoassays have emerged as powerful alternatives, enabling improved accuracy by integrating responses across targets. Such an approach could be particularly valuable when attempting to differentiate infection-induced from vaccine-induced antibody profiles, as it uses subtle variations across multiple immune targets rather than relying on any single antigen^13–17^.

Recently, Jones et al. (2025)^17^ developed and evaluated Mpoxplex, a Luminex-based serological assay specifically designed for mpox diagnostics. This assay distinguishes between infection and MVA-BN vaccine-induced seropositivity, demonstrating high sensitivity and specificity in detecting IgG antibodies against multiple antigens. Their antigen selection was based on prior evidence of immunogenicity and included both MPXV-specific and shared Orthopoxvirus antigens. However, the study had a limited validation period, analyzing samples from a low number of naturally infected and vaccinated individuals (81 and 240 day, respectively). Moreover, given recent findings showing a significant and rapid decline in IgG and neutralizing antibdodies over time post MVA-BN vaccination ^9–11^, it is essential to validate the assay’s diagnostic performance across later time points to ensure its suitability for long-term serosurveillance^9–11,18^. For these reasons, our study builds on the original MpoxPlex assay design—retaining 11 of the initial 12 antigens and incorporating one additional antigen (B16R)—while extending validation to a substantially larger and longitudinally sampled cohort, including follow-up of MVA-BN-vaccinated and MPXV clade IIb convalescent individuals up to two years post exposure.

## Methods

### Study participants and setting

To validate the assay, we used samples collected from an ongoing prospective cohort study of mpox patients and MVA-BN vaccinees conducted at the Institute of Tropical Medicine (ITM), Antwerp, Belgium. The initial results of this study (clinicaltrials.gov registration number NCT05879965) were described previously^9^. Between May 2022 and January 2025, we included 226 individuals, diagnosed with clade IIb MPXV to participate in a long-term follow-up study (**Table 1**). Serum was collected in STT tubes at the moment of diagnosis and during follow-up visits at 15-180 days and 7-10, 15-20, and 23-27 months post-diagnosis. In addition, we included 194 individuals who received the MVA-BN vaccine in 2022 and followed them up during visits at similar timepoints after their second vaccine dose (with 11 individuals also at earlier timepoints). In Belgium, routine smallpox vaccination with a replicating vaccinia virus (VACV)-based vaccine was done until 1976. During the Clade IIb outbreak, the MVA-BN vaccine was used as a primary preventive vaccination (PPV) for individuals at risk from August 2022 onwards. Individuals without a prior childhood vaccination against smallpox received two doses at least 28 days apart, whereas all (except for nine) individuals who had proof of previous smallpox vaccination received only one dose. Participants diagnosed with mpox who also received the MVA-BN vaccine were excluded from analyses. As negative controls, we used samples from 305 patients (182 born after and 123 born before or in 1976) who visited ITM’s clinic before May 2022 and were therefore unexposed to mpox or MVA-BN at the time of sampling.

**Table 1:** Overview of samples in cohorts of MPXV-infected MVA-BN-vaccinated participants, assessed at various months post-infection or vaccination, and negative control from before May 2022.

|  |  | Time post immunizing event<br>(Months) |  |  |  |  |
| --- | --- | --- | --- | --- | --- | --- |
| Age | Mpox/MVA-BN | 0-0.5 | 0.5-6 | 7-10 | 15-20 | 23-27 |
| Born ≤<br>1976 | Infected | 33 | 65 | 27 | 43 | 17 |
|  | vaccinated | 2 | 2 | 72 | 52 | 55 |
| Born<br>>1976 | Infected | 106 | 161 | 74 | 99 | 46 |
|  | Vaccinated | 9 | 5 | 127 | 93 | 85 |

### Luminex multi-analyte IgG immunoassay

Building upon the methodology described by Jones et al^17^, we used commercially available recombinant proteins derived from MPXV and VACV, including VACV A33R, MPXV A35R, VACV B5R, MPXV H3L, MPXV B2R, MPXV A5L, MPXV A27L, MPXV B6R, MPXV E8L, MPXV B22R.64/65, and MPXV B16R (details and suppliers listed in Supplementary Table 1)^19^. Lyophilized proteins were reconstituted in distilled water according to the manufacturers’ guidelines and stored under appropriate conditions until use. Antigen coupling was performed on paramagnetic MAGPLEX COOH-microspheres (Luminex Corporation, Austin, TX) following established protocols^15,20^. Each antigen was coupled at a concentration of 4 µg per 1 × 10^6^ beads. Bovine serum albumin (BSA, Sigma-Aldrich, St. Louis, USA; 3 µg) and tetanus toxoid (EMD Millipore #582231-25UG) were coupled separately to additional bead sets to serve as background controls.

Beads and sera were diluted in PBS containing 1% BSA and 0.05% sodium azide (final serum dilution 1:100) and added to each well, resulting in a total volume of 50 µl per well. Plates were incubated at 37°C on a shaker for 30 minutes, then washed three times with 200 µl/well PBS containing 0.05% Tween 20 and 1%BSA (PBS-TBN). Subsequently, 50 µl of secondary antibody (R-phycoerythrin-conjugated AffiniPure F(ab’)2 fragment of goat anti-human IgG, Jackson ImmunoResearch Laboratories; 1:500) in PBS-TBN was added. The plates were incubated for an additional 30 minutes at 37°C on a shaker set to 600 rpm, protected from light. Following this incubation, plates were washed twice with PBS-TBN, and the beads were finally resuspended in 125 µl PBS-BN.

Data acquisition was performed on a Luminex® 100/200 analyzer, measuring an acquisition volume of 50 µl per well, with double discriminator (DD) gate settings of 5000-25000 and the high photomultiplier tube (PMT) option enabled. Results were reported as crude median fluorescent intensities (MFI). Samples with high background against BSA-coupled beads (defined as greater than four standard deviations [SD] above the mean) or abnormal signals against tetanus toxoid-coated beads (defined as greater or less than four SD from the mean) were retested. Samples consistently exceeding these thresholds after retesting were excluded from further analysis (n=12).

### VACV ELISA and MPXV plaque reduction neutralization test

In addition to the Luminex analyses, all serum samples were tested for IgG binding antibodies using an in-house VACV lysate ELISA, as previously described^10,11^. A random subset of samples was further assessed using an in-house MPXV clade IIb-based plaque reduction neutralization test (PRNT) following established protocols^11^. Binding antibodies against whole *Orthopoxvirus* lysate were measured with ELISA, with 30% endpoint titers ≥ 10 were considered to be positive.

### Statistical analysis

For all analyses, participants were divided into subgroups according to exposure type (infection vs. vaccination) and age. Stratification by age was done to account for prior childhood vaccination. Participants were therefore divided into a group born before or in 1976 and a group born after 1976, the year when routine vaccination with replication-competent VACV was discontinued in Belgium.

We first assessed the predictive value of each antigen individually by generating receiver operating characteristic (ROC) curves and calculating the corresponding Area Under the Curve (AUC) values. For each subgroup, the results obtained from either MPXV-infected individuals or MVA-BN vaccinated individuals were first compared with negative controls from the corresponding age cohort. As a result, for each antigen, four comparisons were generated: (i) MPXV-infected individuals vs. negative controls born after 1976; (ii) MPXV-infected individuals vs. negative controls born in or before 1976; (iii) MVA-BN-vaccinated individuals vs. negative controls born after 1976; and (iv) MVA-BN-vaccinated individuals vs. negative controls born in or before 1976. Because individuals born before or in 1976 are presumed to be vaccinated against smallpox during childhood, comparing MPXV-infected individuals with negative controls born in or before 1976 allowed us to assess the assay’s ability to distinguish natural infection from smallpox-vaccine-induced immunity. Additionally, to directly test if the assay could distinguish natural infection from MVA-BN vaccination, we also compared MPXV-infected individuals vs. MVA-BN-vaccinated individuals in both age cohorts. For each of these comparisons, ROC curves were generated for five distinct post-symptom onset intervals: 0–0.5, 0.5–6, 7–10, 15–20, and 23–27 months.

Subsequently, we explored whether a combined analysis of different antigens could enhance diagnostic accuracy^14^. To this end, we employed a supervised machine learning approach using the random forest algorithm, implemented via the randomForest package in R (version 4.4.2). The dataset was randomly split into a training set (70%) and a validation set (30%) without replacement. The model was trained on the 70% subset using 5,000 trees (ntree = 5000) and three variables per split (mtry = 3), with variable importance estimation enabled. Variable importance was further assessed by calculating the mean decrease in accuracy (MDA) for each antigen, allowing us to identify the five most informative antigens at each time point. We selected maximally five antigens per time point based on preliminary analyses, as this showed no substantial improvement in AUC values when including additional antigens beyond this number. We then evaluated stepwise combinations of these top-ranked antigens to determine the optimal trade-off between predictive performance and cost-efficiency, with the aim of minimizing the number of antigens required to reliably distinguish infection from vaccination. Models were trained on 70% of the data and validated on the remaining 30% using 5000 iterations, with classification performance evaluated using both accuracy metrics and ROC curve analysis. AUC values were computed using the *ROCR* R package^22^ for each post-infection time point. To visualize the gain in predictive power, ROC curves were generated for each incremental antigen combination. To identify which antigens correlated most strongly with neutralizing antibody levels, we correlated the median fluorescence intensity (MFI) values of the different Luminex antigens to PRNT_50_ values using the *corrplot* package in R. For this analysis, we excluded MVA-BN vaccinated individuals.

Finally, we investigated whether our multiplex antigen panel could also provide information on the timing of MPXV infection based on antigen-specific IgG profiles. Using the same random forest classifier and settings, we trained a model to distinguish between samples collected at five time points post-infection (0–0.5, 0.5–6, 7–10, 15–20, and 23–27 months) based on their Luminex antibody signatures. The classification performance was evaluated using confusion matrices and variable importance measures.

### Ethical consideration

All participants in the prospective mpox and vaccine cohorts provided written informed consent prior to any study procedures. Individuals included in the retrospective arm of the mpox cohort were enrolled based on our clinic’s standard policy, provided they had not opted out of the use of their data and samples for research purposes. Ethical approvals were granted by the Institutional Review Board of the Institute of Tropical Medicine (ITM; references 1628/22, 1641/22 and 34/2023) and the Ethics Committee of the University Hospital of Antwerp (UZA; references 3797 and 4981). The study was conducted in accordance with the Declaration of Helsinki and adhered to the latest Good Clinical Practice (GCP) guidelines.

## Results

### Accuracy of individual antigens

IgG levels against the different antigens decreased over the two-year study period, both in mpox-infected individuals (**Fig. 1**) and MVA-BN vaccine recipients (**Fig. 2**). These results are consistent with our previous findings, which indicated a gradual decline in neutralizing and binding antibody titers over time in this cohort^9^. In addition, baseline (pre-exposure) antibody titers were higher among participants born before or in 1976. Correspondingly, ROC analyses revealed that the predictive performance of individual antigens varied depending on both the time since symptom onset and childhood vaccination status. Different patterns emerged in the four subgroups which are based on type of exposure (natural infection vs vaccination) and on childhood vaccination (born before or in 1976 vs born after 1976).

**Fig 1:**
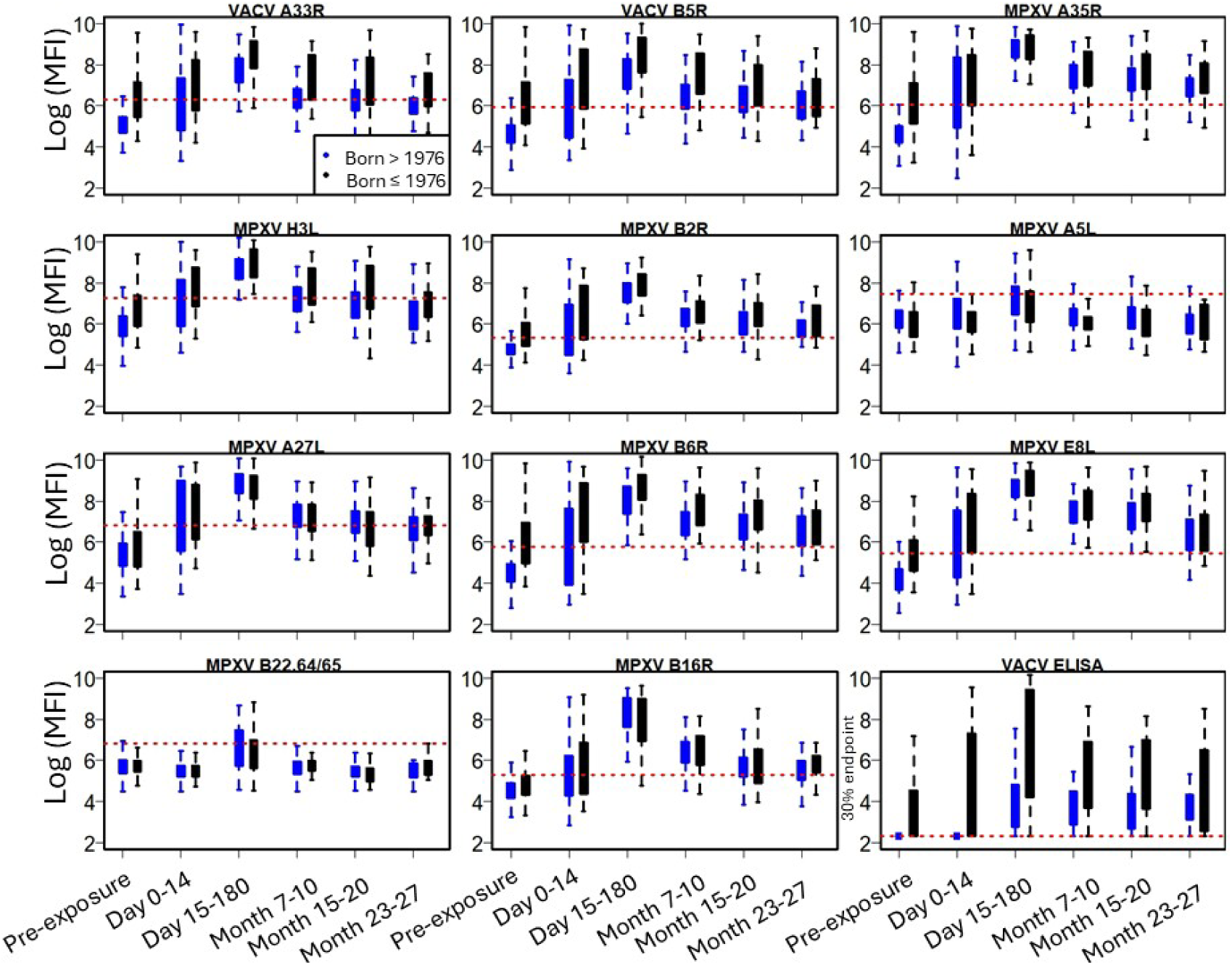
Distribution of IgG titers in MPXV-infected individuals over time since symptom onset, as measured by the Luminex assay (log2 MFI values) and VACV ELISA (30% endpoint titers). The red horizontal line represents the 95% specificity cutoff, established using 196 pre-2022 samples from naive individuals born after 1976. Colors reflect participants’ birth year, inferring prior smallpox vaccination status (black = born before or in 1976; blue = born after 1976). E8 and VACV ELISA results are also presented in Van Dijck et al^9^

**Fig 2:**
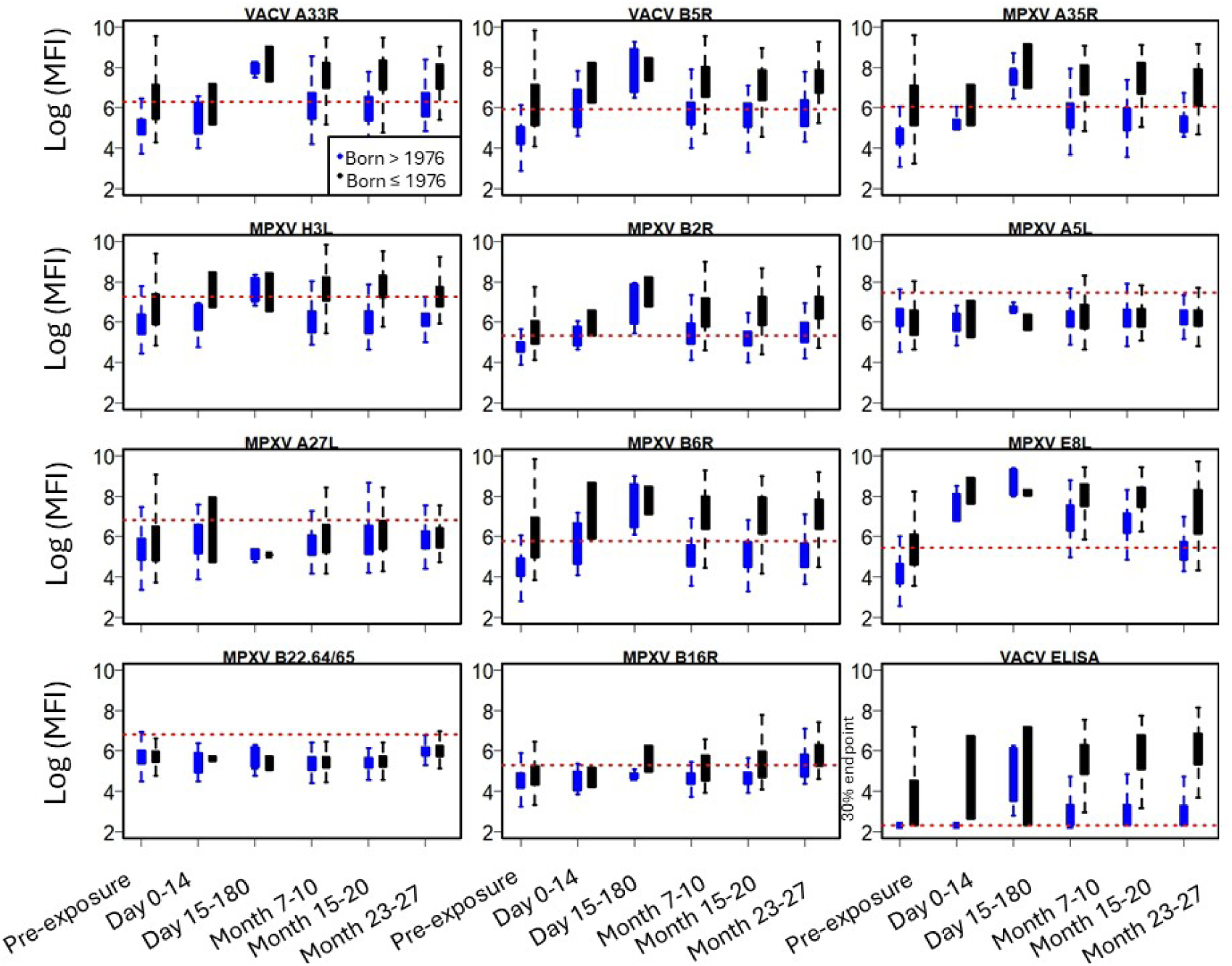
Distribution of IgG titers in MVA-BN vaccinated individuals over time since symptom onset, as measured by the Luminex assay (log2 MFI values) and VACV ELISA (30% endpoint titers). The red horizontal line represents the 95% specificity cutoff, established using 196 pre-2022 samples from naive individuals born after 1976. Colors reflect participants’ birth year, inferring prior smallpox vaccination status (black = born before or in 1976; blue = born after 1976). E8 and VACV ELISA results are also presented in Van Dijck et al^9^.

In MPXV-infected individuals who did not receive childhood vaccination (born after 1976), most antigens—excluding B22R.64/65, H3L, and A5L—reliably detected IgG antibodies from two weeks up to 20 months post infection. When compared with negative controls from the same age cohort, these antigens yielded area under the curve (AUC) values exceeding 0.85 and demonstrated sensitivities above 80% at 95% specificity (**Fig. 3; Supplementary Table 1**). However, the diagnostic performance declined at later time points, as the AUC and sensitivity values declined for most antigens. Nevertheless, MPXV antigens A35R, B2R, B6R, and E8L maintained excellent diagnostic accuracy, with AUC values consistently above 0.94 and sensitivities exceeding 0.80 even two years post infection, underscoring their potential for reliable long-term serological surveillance. In contrast, antigens such as A27L and H3L showed a more rapid decline in performance over time. As such, they might be better suited for identifying recent infections or re-infections.

**Fig 3:**
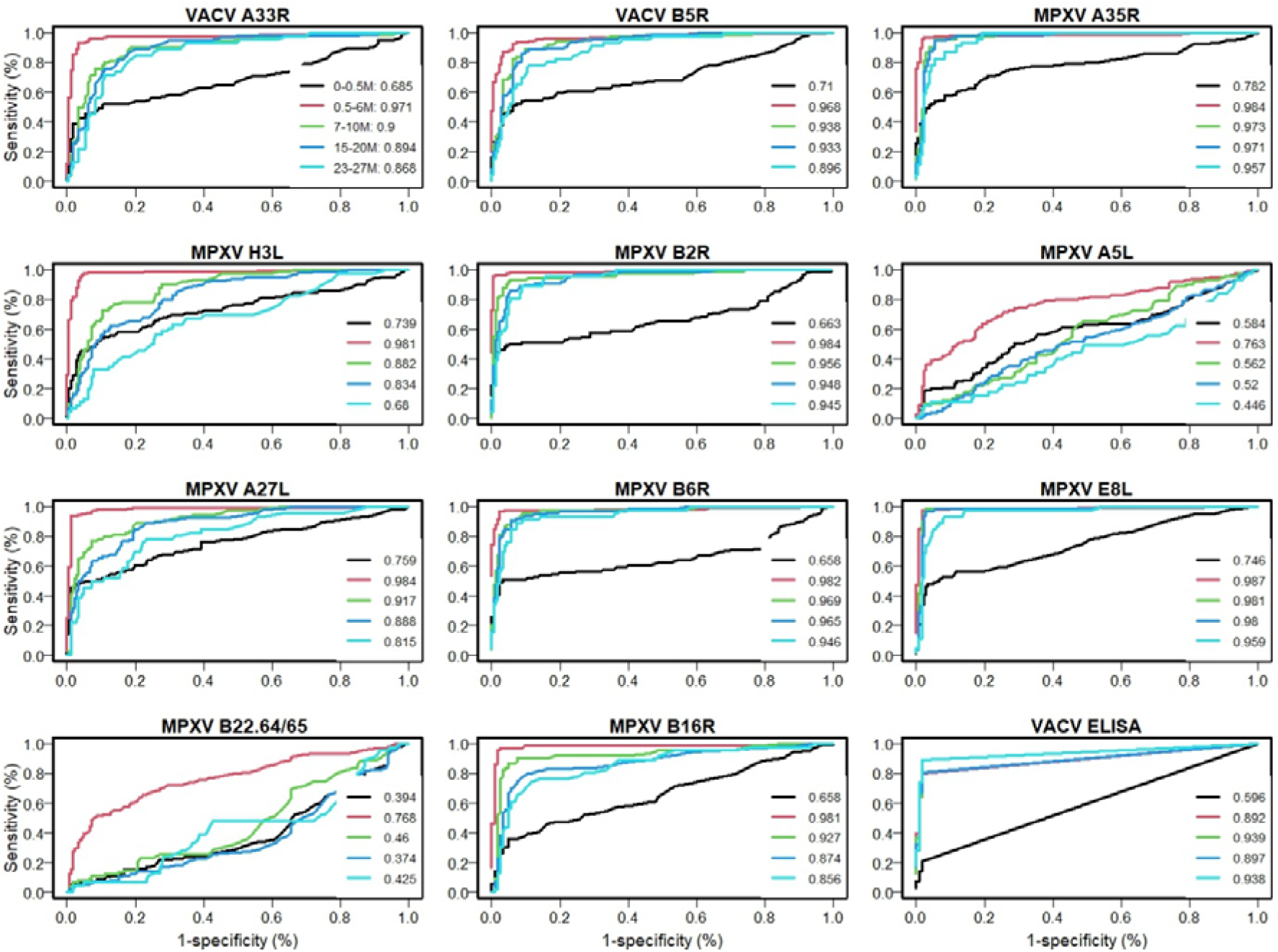
Receiver operator characteristics for different antigens of the IgG Luminex assay and VACV ELISA, comparing MPXV infected with negative control samples from individuals born after 1976 (smallpox vaccine-naive). Different colours represent different time periods post symptom onset (at 0-0.5, 10.5-6, 7-10, 15-20 and 23-27 months). Area under the curve (AUC) values are presented in the legend.

In MPXV-infected individuals with prior smallpox vaccination during childhood (born before or in 1976), the antigens showed reduced discriminatory power for detecting past mpox infection, when compared with negative controls from the same age group (**Fig. 4; Supplementary Table 1**). Although most antigens performed reasonably well up to six months after infection, their AUC values dropped with sensitivity below 0.6 at later time points. The E8L antigen was a notable exception, with AUC values of 0.91 and a sensitivity of 0.82 up to 10 months after infection.

**Fig 4:**
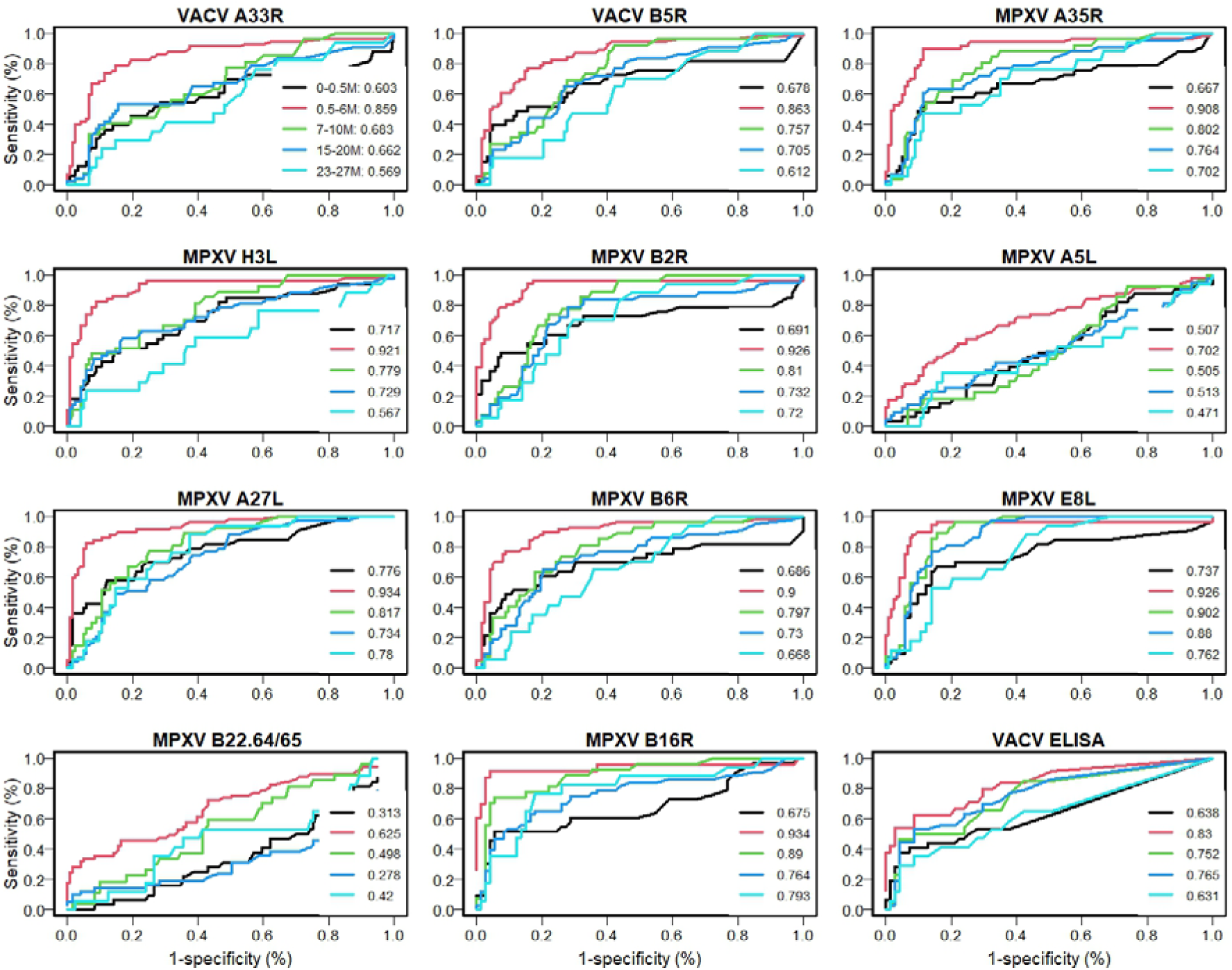
Receiver operator characteristics for different antigens of the IgG Luminex assay and VACV ELISA, comparing MPXV infected with negative control samples from individuals born in or before 1976 (smallpox vaccines). Different colours represent different time periods post symptom onset (at 0-0.5, 0.5-6, 7-10, 15-20 and 23-27 months). Area under the curve (AUC) values are presented in the legend.

In MVA-BN–vaccinated individuals without prior smallpox vaccination (born after 1976), IgG titers for nearly all antigens declined below the assay’s 95% specificity cutoff by seven months post-vaccination (**Fig 2**). This decline was accompanied by a drop in AUC values below 0.85 and corresponding sensitivities falling below 0.60, indicating limited diagnostic utility beyond this time point. (**Supplementary Fig. 1; Supplementary Table 1**). Again, the E8L antigen stood out, maintaining AUC values of 0.97 and 0.88 at 15-20 and 23-27 months post-vaccination, respectively. Similarly, in MVA-BN–vaccinated individuals born in or before 1976, and therefore likely vaccinated during childhood, the assay showed a low accuracy in distinguishing vaccinated individuals with negative controls for the same age cohort. Except for E8L, AUC values dropped below 0.85 at months 7-10 with sensitivities below 0.60 for all other antigens (**Supplementary Fig. 2; Supplementary Table 2**). Finally, when we analyzed the assay’s ability to distinguish between natural infection and vaccination by comparing MPXV-infected individuals and MVA-BN vaccinees in the different birth cohorts, we found AUC values consistently at or below 0.85, while sensitivity fell below 0.5 (**Supplementary Figs 3 and 4**).

### Correlations between antigens, ELISA and neutralizing antibodies

We observed strong positive correlations (pairwise Spearman’s ρ values > 0.7) between most individual antigens and neutralizing antibody titers in mpox infected individuals (**Fig 5**). Separately, we observed moderate-to-strong correlations between antibody titers against the different antigens included in the assay (**Fig. 5**), with pairwise Spearman’s ρ values typically ranging between 0.60–0.8. This indicates a broadly reactive immune response without clear dominance by a single antigen. Notably, MPXV A5L and B22R.64/65 were exceptions, showing weak or no correlation with neutralization titers or other antigens in the assay (pairwise Spearman’s ρ values < 0.6). These findings suggest that antibody responses to most antigens are positively associated with neutralizing titers, whereas responses to MPXV A5L and B22R.64/65 appear to follow a different pattern, potentially reflecting distinct or non-neutralizing immune activity.

**Fig 5:**
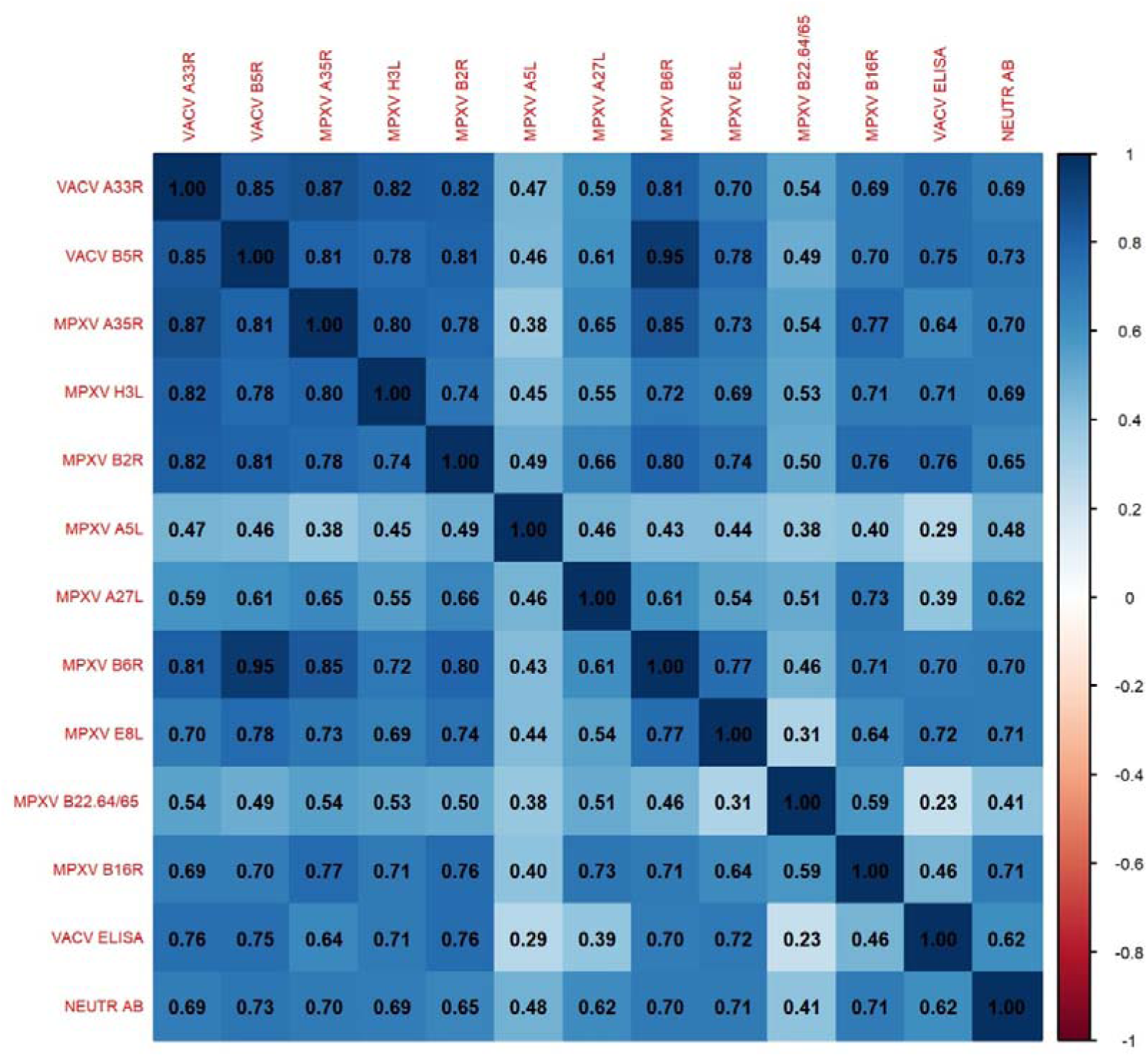
Correlation matrix between the different antigens of the Luminex, the VACV ELISA and neutralization assay, using titers from MPXV infected individuals only. Blue indicates high positive correlations between variables, white represents no correlations and red negative correlations.

### Accuracy of multiplexing antigens

To explore if combinations of antigens could improve the accuracy of our assay, we applied a random forest algorithm to evaluate the relative importance of each antigen at different time points, using the mean decrease in accuracy (MDA) as a measure of contribution (**S Fig. 5**). Consistent with the above described analysis of individuals antigens, MPXV E8L, B5R, and B2R and B6R showed the highest impact on classification accuracy two years after natural infection. By combining the top five antigens, the predictive performance of the assay to diagnose previous mpox infection from negative controls improved at later time points both among individuals born after (**Fig 6**) or before 1976 (**S Fig 6**), with an increase in AUC of 10 and 18, respectively. Specifically, 23 months post infection, a panel comprising B6R, B5R, and E8L was most effective in predicting infection in individuals born after 1976 (AUC = 0.98, sensitivity = 0.8) (**Fig 6**). For the same time period, a panel of B6R, B5R, A35R was most effective in predicting infection in individuals born before or in 1976 (AUC = 0.73, sensitivity = 0.35) (**S. Fig 6**).

**Fig 6:**
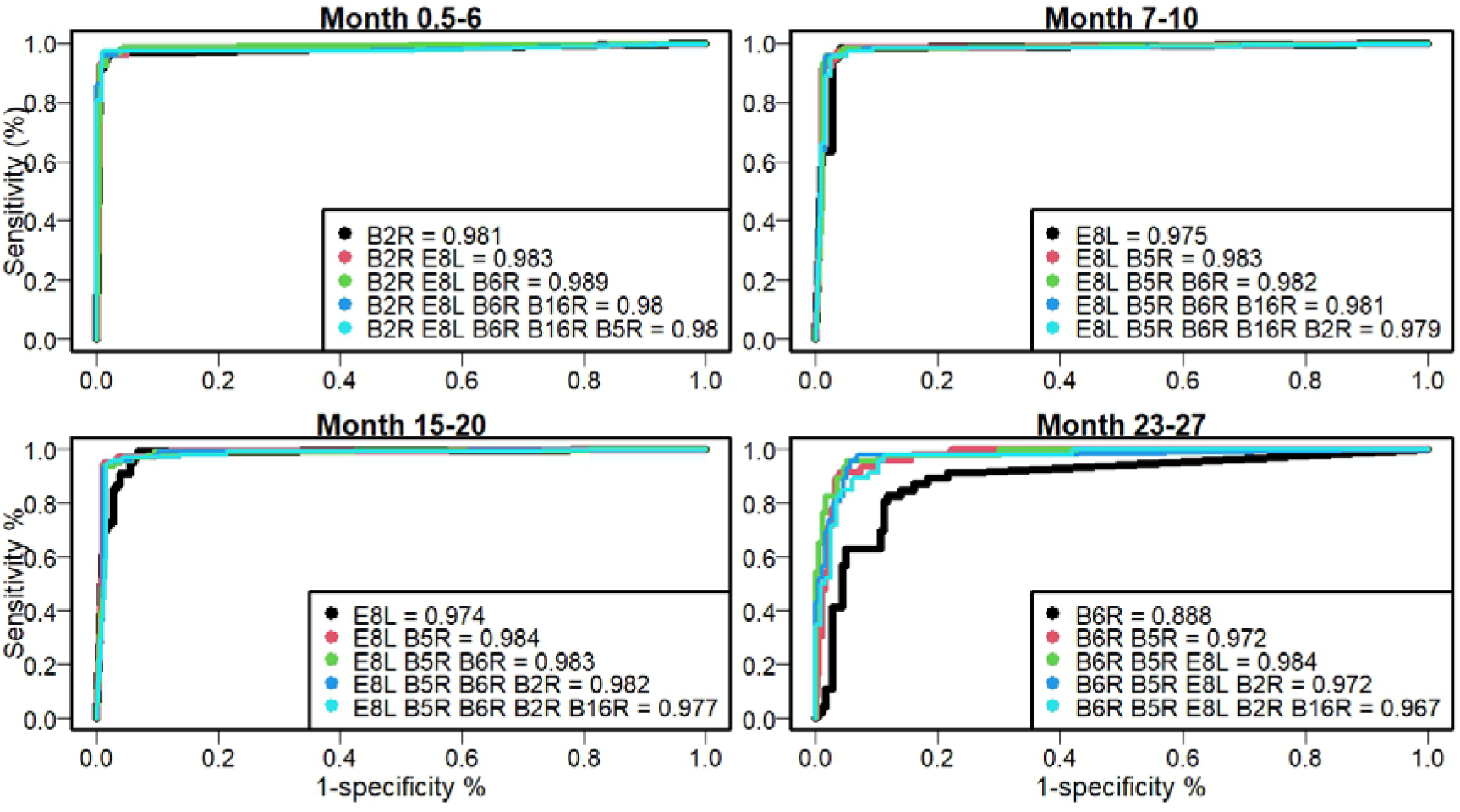
Receiver operator characteristics for different combinations of antigens for the IgG Luminex assay on samples, comparing MPXV infected with negative control samples from individuals born after 1976 (smallpox vaccine-naïve) at different time periods post symptom onset (at 0.5-6, 7-10, 15-20 and 23-27 months). Different colours represent different combinations of antigens. Area under the curve (AUC) values are presented in the legend.

### Temporary signal predictions

To explore whether based on antigen-specific IgG profiles it is possible to estimate temporary signals (i.e. the time since natural infection), we trained a random forest classifier using all antigens in the assay to assign samples to predefined post-infection time intervals and evaluated its performance using a confusion matrix (**Fig 7**). Among individuals born after 1976, the model correctly classified 79% of samples collected within the first six months post-infection, with antigens E8L, A27L, and B16R emerging as the most informative features based on variable importance metrics (**S Fig 7**). However, classification accuracy declined substantially beyond this early window. For samples collected ≥20 months post-infection, only 20% were correctly classified, and there was increasing overlap across intermediate time intervals. This indicates that while the model can reliably distinguish recent infections (<6 months) based on IgG profiles, its ability to resolve more precise timing diminishes with increasing time since infection. As such, the assay provides useful temporal resolution primarily for identifying recent versus older infections, rather than pinpointing the exact time since exposure.

**Fig 7:**
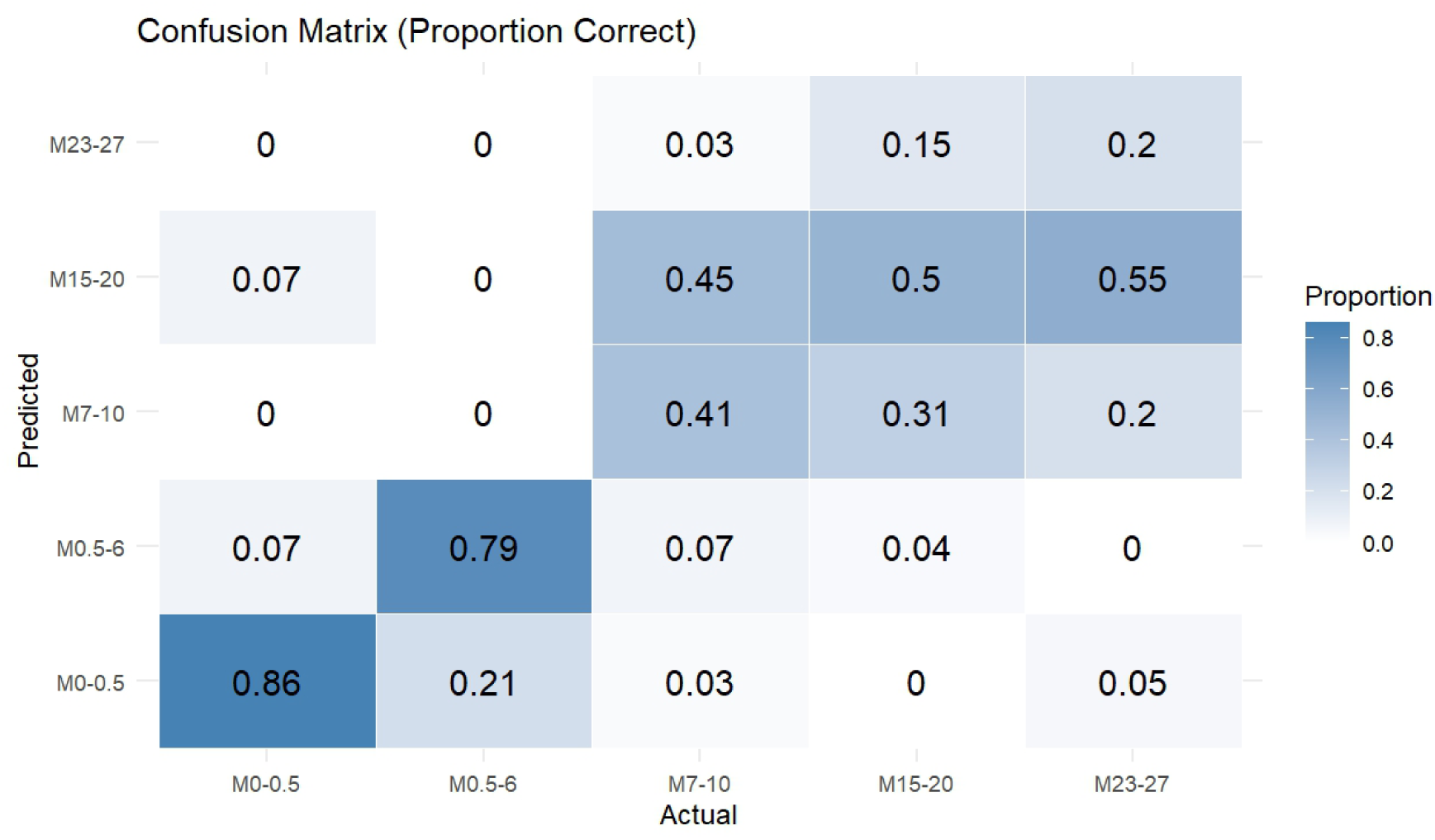
Confusion matrix showing the classification performance of the model in predicting month since MPXV infection based on IgG profiles, when comparing MPXV infected with negative control samples from individuals born after 1976 (smallpox vaccine-naïve).

## Discussion

In this study, we assessed the performance of an 11-antigen multiplex immunoassay, called Mpoxplex, for detecting IgG antibodies up to two years after natural mpox infection or vaccination. The assay showed strong long-term discriminatory capacity for diagnosing previous mpox infections, particularly in individuals without childhood vaccination. In contrast, the assay’s performance in detecting previous vaccination with MVA-BN decreased substantially from six months post vaccination onwards.

This is consistent with recent findings from our group and others, which show that MVA-BN– induced antibody responses decline substantially over time^9–11^. By one year post-vaccination, most individuals had neutralization titers approaching or falling below the detection threshold^10^. Our results confirm these observations, with IgG levels for most Luminex antigens falling below the assay cutoff as early as seven months post-vaccination. Notably, Jones et al. (2025)^17^ reported more persistent IgG titers at seven months post-vaccination for several antigens, including B6R, A33R, A35R, and E8L. However, their study was based on a small sample size (n=8). In our cohort, the E8L antigen, associated with surface attachment and viral entry^19^, stood out as an exception, with antibody titers remaining above the assay cutoff for up to 20 months post-vaccination. Given its sustained performance, E8L could likely serve as a reliable singleplex marker for long-term detection of vaccine-induced immunity.

Among MPXV-infected individuals born after 1976, several MPXV antigens (A35R, B2R, B6R, and E8L) demonstrated strong predictive power (ROC AUC>0.94) in singleplex format, up to two years post-infection. In contrast, the assay performed considerably less well in differentiating MPXV infection from smallpox or MVA-BN vaccinated individuals. In these groups, multiplexing antigens improved performance during early time points with sensitivity increasing from 51% to 83% (at 95% specificity) when combining A27L, H3L and B16R. These results from earlier time points are in line with results from other studies^15,23^. However, at later time points, the discriminatory power diminished substantially, even when combining antigens (AUC < 0.75 up to 2 years). So far, no serological test has been able to reliably distinguish MPXV-infection with vaccinia-based vaccination at these later time points, which poses a challenge for population-based surveillance, especially when done in age groups that have received smallpox vaccination during childhood.

We further tested whether specific antigens on the Luminex assay could serve as a proxy for neutralization, which is the gold standard for assessing functional antibody responses^11,24^. Neutralizing antibodies are widely recognized as a key correlate of protection against many viral infections, potentially including mpox^10,25^. In our analysis, we observed strong positive correlations between most antigens tested and neutralizing antibody levels, indicating robust concordance between binding and neutralizing immune responses, as observed in other viral diseases^26^. This suggests that high-throughput Luminex-based assays, which are more cost-effective and less reliant on specialized biosafety infrastructure than neutralization tests, may serve as practical surrogates for neutralization in sero-epidemiological studies. Such an approach would be especially valuable in low-resource settings like the Democratic Republic of Congo, where mpox is most prevalent and laboratory capacity is often limited^4,27^.

Finally, we investigated the potential of using multiple antigens to retrospectively estimate the timing of MPXV infections, as this enables better estimates of waning immunity and general transmission dynamics^28^. While mpox often presents with distinctive symptoms that could, in theory, facilitate the determination of infection timing, some cases may be subclinical or asymptomatic^29,30^. Serological data represent a promising alternative for inferring time since infection and are particularly powerful when based on experimental infection data, which help calibrate these models^28,31^. Our time-calibrated model demonstrated modest classification accuracy within the first three months post-infection, with about 79% of samples correctly classified, but accuracy declined as the infection became more distant in time. This indicates that while antigen-specific IgG profiles carry a strong temporal signal shortly after infection, these signals degrade over time to less than 50%, limiting precise timing estimates for older infections. Nonetheless, the ability to distinguish recent from past infections holds considerable potential for retrospective epidemiological studies.

A limitation of our study is the absence of external validation of our random forest classifier using an independent cohort, which may limit the generalizability of the multiplexing to other populations or sampling conditions. Additionally, all mpox cases included in our cohort were predominantly symptomatic. As a result, we cannot exclude the possibility that subclinical or asymptomatic infections could result in lower antibody levels and, consequently, reduced assay sensitivity in such cases.

In conclusion, our study demonstrated that a subset of antigens from the MpoxPlex assay are robust and reliable for long-term serological detection of MPXV infection, maintaining strong diagnostic performance for up to two years post-infection. These antigens reliably distinguished MPXV infection from uninfected individuals, but only among non-vaccinated individuals. We showed strong correlation with neutralizing antibody levels, and that multiplexing antigens furthers improves classification accuracy. Although predicting the exact timing of past infection remains challenging, our findings support the use of multiplexing key antigens to differentiate recent from older exposures. Taken together, these results highlight the assay’s potential for use in serosurveillance in both endemic and non-endemic settings, although previous vaccination status should be carefully considered.

## Data Availability

The study de-identified dataset is available from the corresponding author upon reasonable request.

## Contributors

Conceptualized the study and wrote the protocol: JM, CVD, NBR, LL. Data collection: SB,JV, FvR, CC, EB, NdC, BS, LV, AvH, LM; Human sampling design: BS, OG; Conducted sample processing and analysis: EW, LZ, SJ, BH, AO, JLY, DTH, WTL,; Conducted data analysis: CVD, NBR, JM, CK, AT, LMZ. Data management: CVD, NBR, JM, LMZ. Secured funding: LL, JM, KA. Supervision study proceedings: LL, KA,JM, RDdV, WA,KV, MvE, AM, PS. Supervised laboratory procedures: JM, KKA, WA, RDdV, MVE, KV. Writing of the initial manuscript: JM. Funding acquisition: JM, LL, LMZ, RDdV. Reviewed the manuscript: all authors. All authors contributed to review and approved the final version of the manuscript.

## Data sharing

The study’s de-identified dataset is available from the corresponding author upon reasonable request.

## Acknowledgements

During the preparation of this work, the authors used ChatGPT-4o as an editing tool (prompt: “improve grammar and spelling”). After using this tool, the authors reviewed and edited the content as needed and take full responsibility for the content of the publication.

## Supplementary tables

**S table 1:** Overview of orthopoxvirus antigens included in the MpoxPlex assay, along with their viral origin and commercial suppliers.

|  |  |  |
| --- | --- | --- |
| VACV<br>A33R | Vaccinia Virus (WR) A33RR Protein with C-terminal Histidine Tag, Recombinant from baculovirus | Beiresources |
| MPXV<br>A35R | Recombinant Monkeypox Virus EEV Envelope Glycoprotein, A35RR | RayBiotech |
| VACV<br>B5R | Vaccinia Virus (WR) B5RR Protein with N-terminal Histidine Tag, Recombinant from baculovirus | Beiresources |
| MPXV<br>B2R | Monkeypox Virus Hemagglutinin (MPXV B2RR) Protein | Abbexa |
| MPXV<br>H3L | Recombinant Monkeypox Virus Envelope Protein H3LL | RayBiotech |
| MPXV<br>A5L | Monkeypox virus gp115 / 39kDa protein, C-terminal His-tag | The Native Antigen Company |
| MPXV<br>A27L | Recombinant Monkeypox virus/MPXV A27LL Protein | Proteogenix |
| MPXV<br>B6R | Monkeypox Virus (MPXV) B6RR Protein (His Tag) | Sino Biological |
| MPXV<br>E8L | Recombinant Monkeypox virus/MPXV E8L Protein | Proteogenix |
| MPXV<br>B22R | B22R.64/65 (peptide) | Eurogentec |
| MPXV<br>B16R | Recombinant Monkeypox Virus IFN-alpha/beta-receptor-like secreted Glycoprotein B16RR | RayBiotech |

**S table 2:** Area Under the Curve (AUC) values for different monkeypox or vaccinia antigens after mpox infection or MVA-BN vaccination at different months post infection/vaccination, compared with negative control samples from individuals obtained before the mpox epidemic in Belgium.

|  |  | Mpox infection |  |  |  |  | MVA-BN |  |  |
| --- | --- | --- | --- | --- | --- | --- | --- | --- | --- |
|  | Months | 0-0.5 | 0.5-6 | 7-10 | 15-20 | 23-27 | 7-10 | 15-20 | 23-27 |
| Born After 1976 | VACV A33R | 0,69 | 0,97 | 0,9 | 0,89 | 0,87 | 0,83 | 0,79 | 0,87 |
|  | MPXV A35R | 0,79 | 0,98 | 0,97 | 0,97 | 0,96 | 0,79 | 0,76 | 0,75 |
|  | VACv B5R | 0,71 | 0,97 | 0,93 | 0,93 | 0,89 | 0,84 | 0,82 | 0,84 |
|  | MPXV H3L | 0,73 | 0,98 | 0,87 | 0,83 | 0,68 | 0,53 | 0,54 | 0,64 |
|  | MPXV B2R | 0,67 | 0,98 | 0,94 | 0,94 | 0,94 | 0,78 | 0,75 | 0,81 |
|  | MPXV A5L | 0,58 | 0,74 | 0,57 | 0,51 | 0,44 | 0,47 | 0,53 | 0,52 |
|  | MPXV A27L | 0,76 | 0,98 | 0,90 | 0,88 | 0,81 | 0,57 | 0,64 | 0,66 |
|  | MPXV B6R | 0,67 | 0,98 | 0,95 | 0,95 | 0,94 | 0,71 | 0,68 | 0,73 |
|  | MPXV E8L | 0,75 | 0,99 | 0,98 | 0,98 | 0,96 | 0,98 | 0,97 | 0,87 |
|  | MPXV B22R | 0,41 | 0,77 | 0,46 | 0,36 | 0,43 | 0,33 | 0,34 | 0,67 |
|  | MPXV B16R | 0,65 | 0,98 | 0,91 | 0,87 | 0,86 | 0,58 | 0,59 | 0,82 |
|  | VACV ELISA | 0,59 | 0,9 | 0,93 | 0,89 | 0,94 | 0,72 | 0,78 | 0,79 |
| Born before 1976 | VACV A33R | 0,60 | 0,86 | 0,68 | 0,66 | 0,57 | 0,76 | 0,77 | 0,77 |
|  | MPXV A35R | 0,67 | 0,91 | 0,81 | 0,76 | 0,71 | 0,74 | 0,76 | 0,69 |
|  | VACv B5R | 0,68 | 0,86 | 0,76 | 0,72 | 0,61 | 0,74 | 0,75 | 0,78 |
|  | MPXV H3L | 0,71 | 0,92 | 0,78 | 0,73 | 0,56 | 0,74 | 0,77 | 0,71 |
|  | MPXV B2R | 0,69 | 0,92 | 0,82 | 0,72 | 0,72 | 0,75 | 0,76 | 0,81 |
|  | MPXV A5L | 0,51 | 0,71 | 0,52 | 0,51 | 0,47 | 0,58 | 0,54 | 0,57 |
|  | MPXV A27L | 0,78 | 0,93 | 0,82 | 0,73 | 0,78 | 0,57 | 0,62 | 0,61 |
|  | MPXV B6R | 0,69 | 0,91 | 0,81 | 0,74 | 0,68 | 0,71 | 0,71 | 0,71 |
|  | MPXV E8L | 0,74 | 0,92 | 0,91 | 0,88 | 0,76 | 0,91 | 0,91 | 0,83 |
|  | MPXV B22R | 0,3 | 0,61 | 0,47 | 0,26 | 0,42 | 0,31 | 0,34 | 0,63 |
|  | MPXV B16R | 0,68 | 0,94 | 0,89 | 0,78 | 0,79 | 0,69 | 0,71 | 0,85 |
|  | VACV ELISA | 0,62 | 0,84 | 0,76 | 0,77 | 0,63 | 0,82 | 0,84 | 0,88 |

## Supplementary figures

**S Fig 1:**
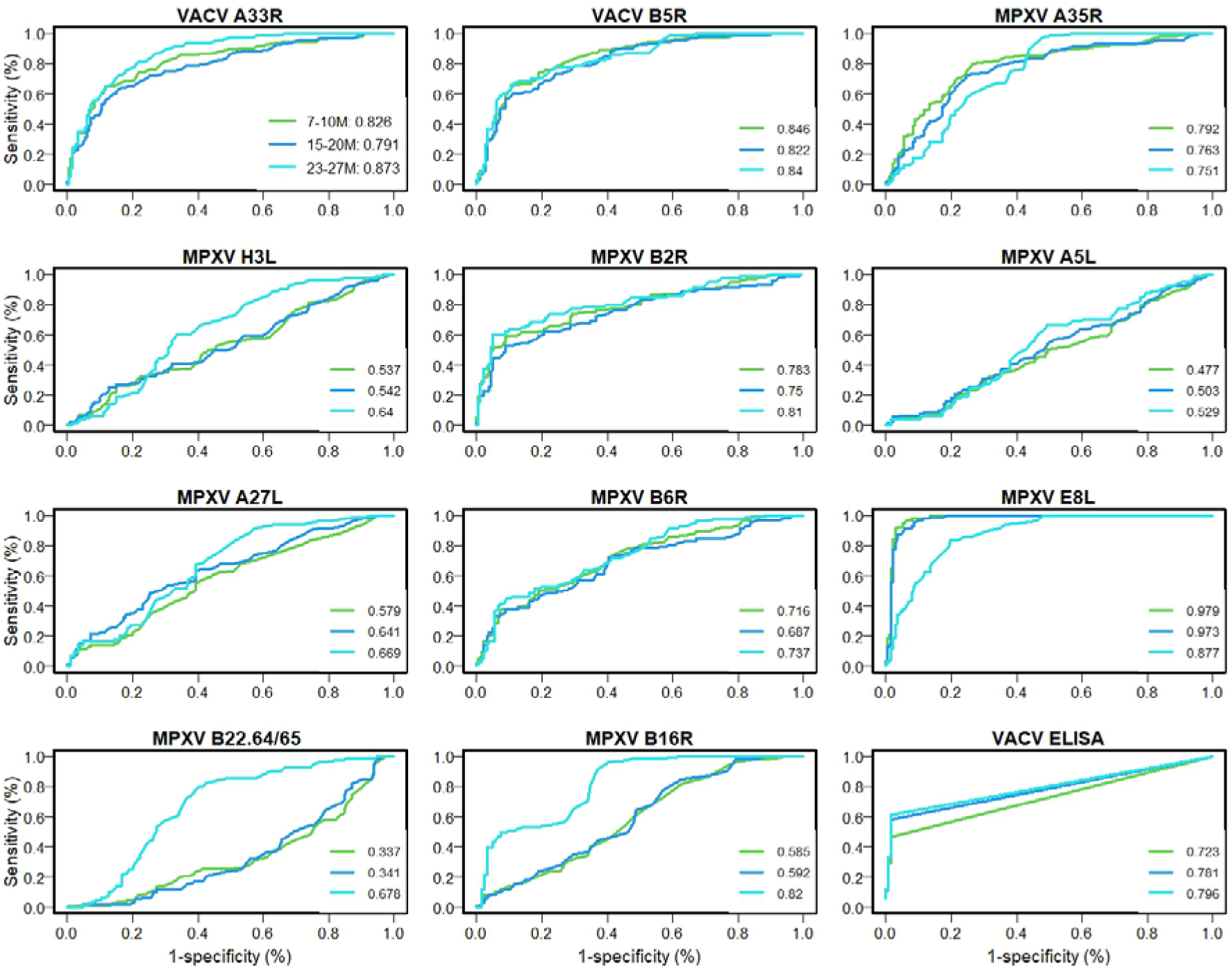
Receiver operator characteristics for different antigens of the IgG Luminex assay and VACV ELISA, comparing MVA-BN vaccinated vs. negative control samples from individuals born after 1976 (smallpox vaccine-naive). Different colours represent different time periods post symptom onset (at 7-10, 15-20 and 23-27 months). Area under the curve (AUC) values also presented in the legend.

**S Fig 2:**
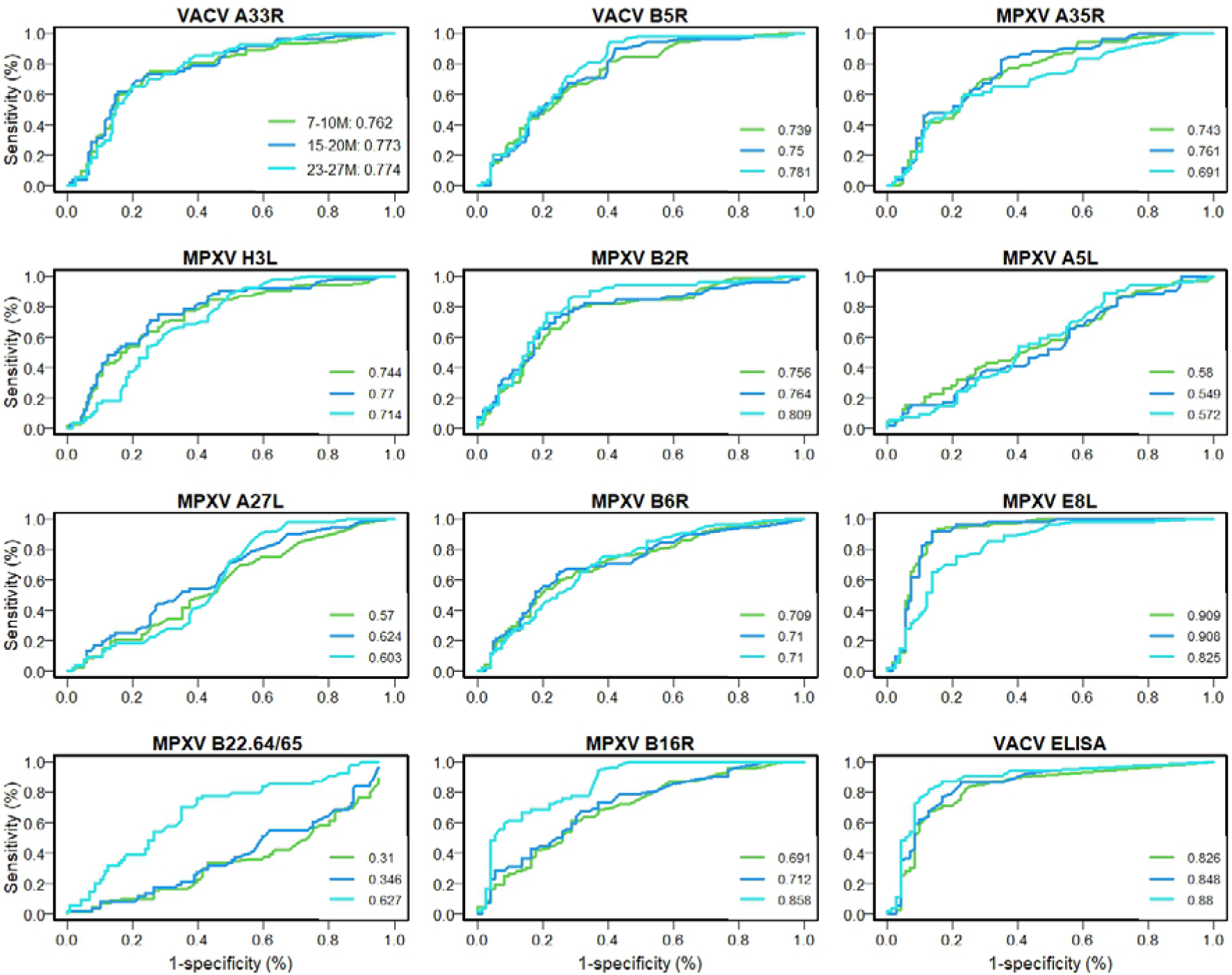
Receiver operator characteristics for different antigens of the IgG Luminex assay and Vaccinia ELISA, comparing MVA-BN vaccinated vs. negative control samples from individuals born in or before 1976 (smallpox vaccine-naive). Different colours represent different time periods post symptom onset (at 7-10, 15-20 and 23-27 months). Area under the curve (AUC) values are presented in the legend.

**S Fig 3:**
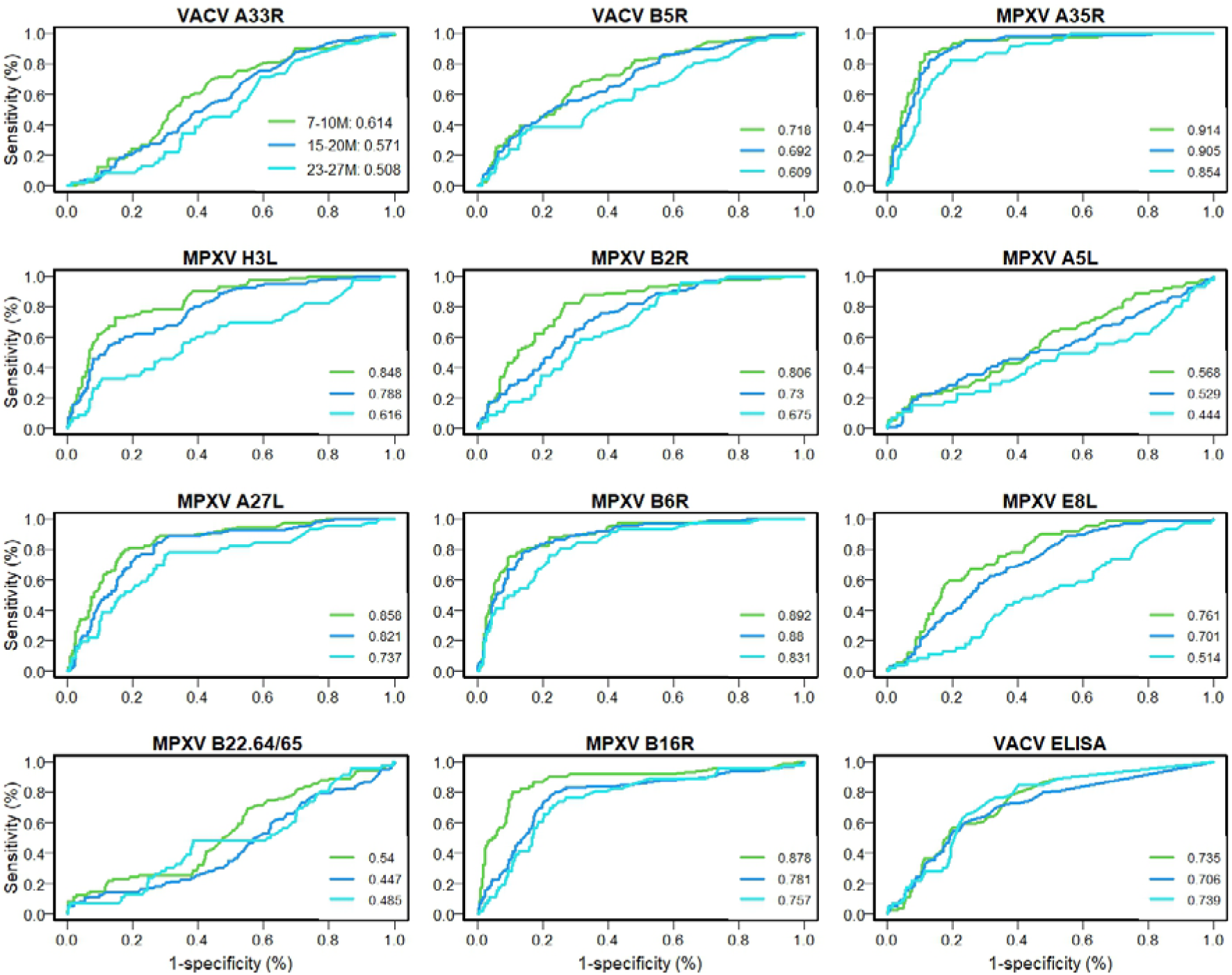
Receiver operator characteristics for different antigens of the IgG Luminex assay and Vaccinia ELISA, comparing MVA-BN vaccinated vs. mpox infection samples from individuals born after 1976 (smallpox vaccine-naive). Different colours represent different time periods post symptom onset (at 7-10, 15-20 and 23-27 months). Area under the curve (AUC) values are presented in the legend.

**S Fig 4:**
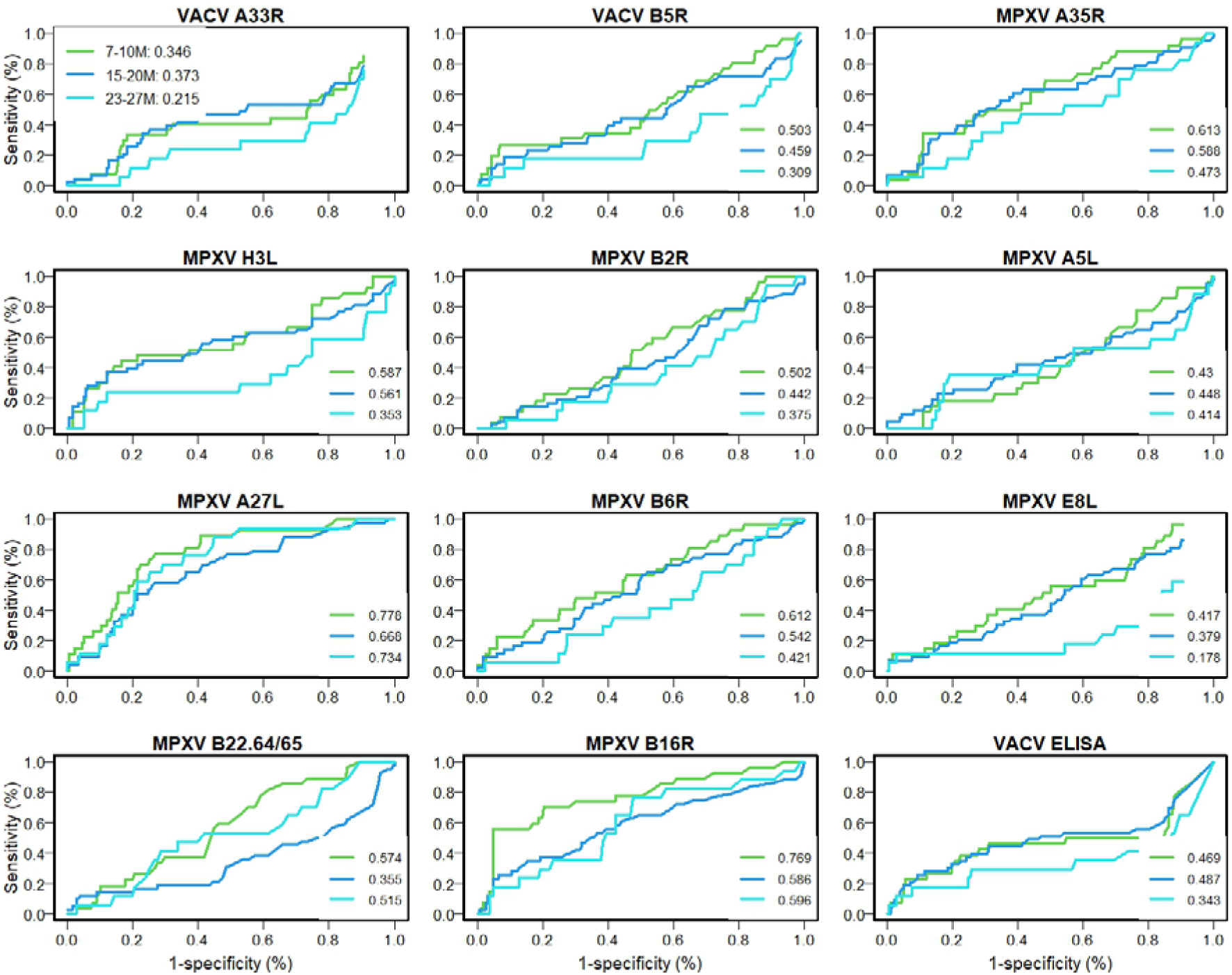
Receiver operator characteristics for different antigens of the IgG Luminex assay and Vaccinia ELISA, comparing MVA-BN vaccinated vs. mpox infected samples from individuals born before or in 1976 (smallpox vaccine-naive). Different colours represent different time periods post symptom onset (at 7-10, 15-20 and 23-27 months). Area under the curve (AUC) values are presented in the legend.

**S Fig 5:**
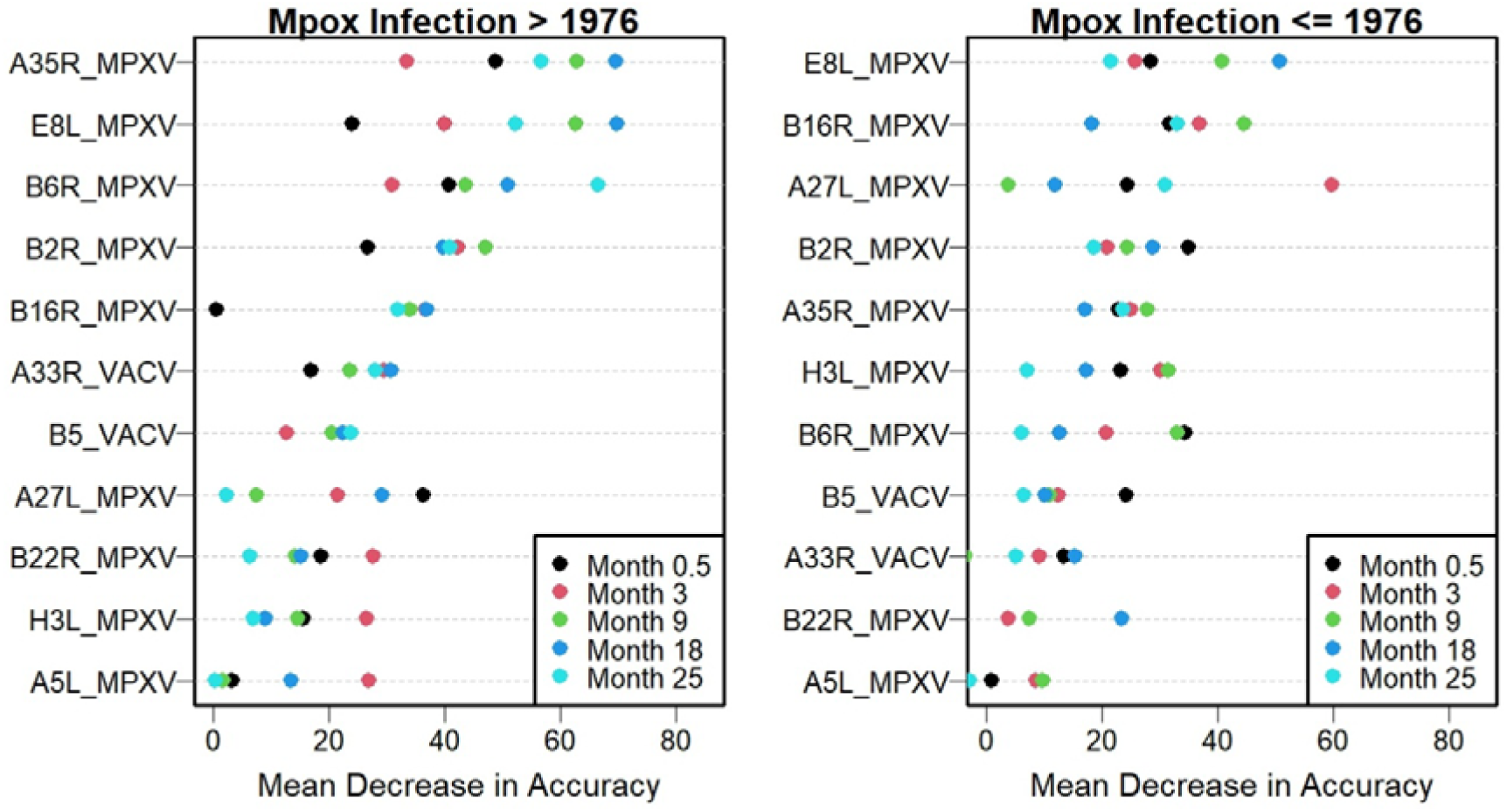
Mean decrease in accuracy (MDA) for different antigens in the IgG Luminex assay for different periods post symptom onset (at 0-0.5, 1-6, 7-10, 15-20 and 23-27 months). **Left**: MDA when comparing MPXV infected vs. negative control samples from individuals born after 1976 (smallpox vaccine-naïve); **right**: MDA when comparing MPXV infected vs. negative control samples from individuals born in or before 1976 (smallpox vaccine-naïve)

**S Fig 6.**
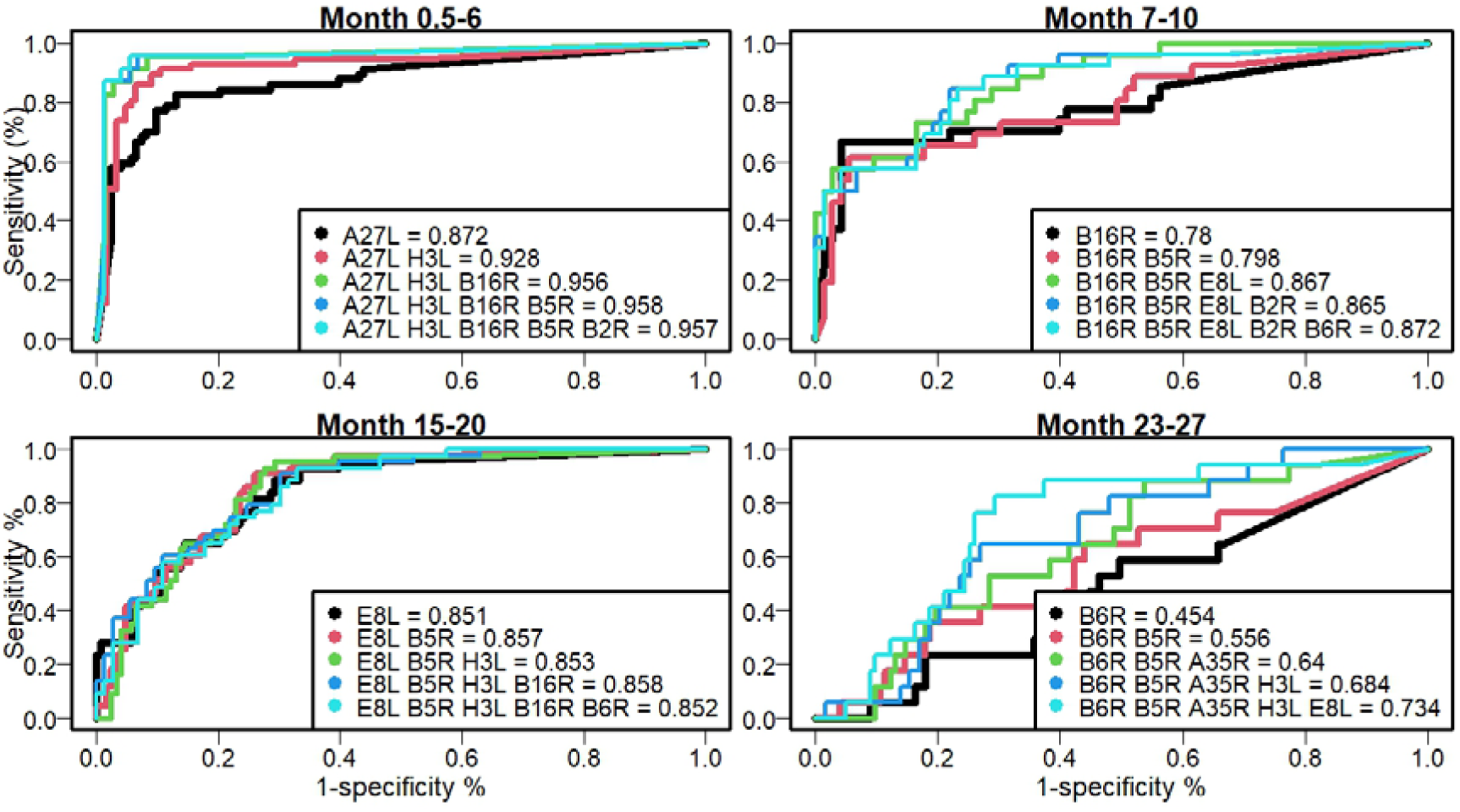
Receiver operator characteristics for different antigens of the IgG Luminex assay, comparing mpox infected vs. negative control samples from individuals born in or before 1976 (smallpox vaccine-naive) at different time periods post symptom onset (at 0.5-6, 7-10, 15-20 and 23-27 months). Different colours represent different combinations of antigens. The area under the curve values (AUC) is presented in the legend.

**S Fig 7:**
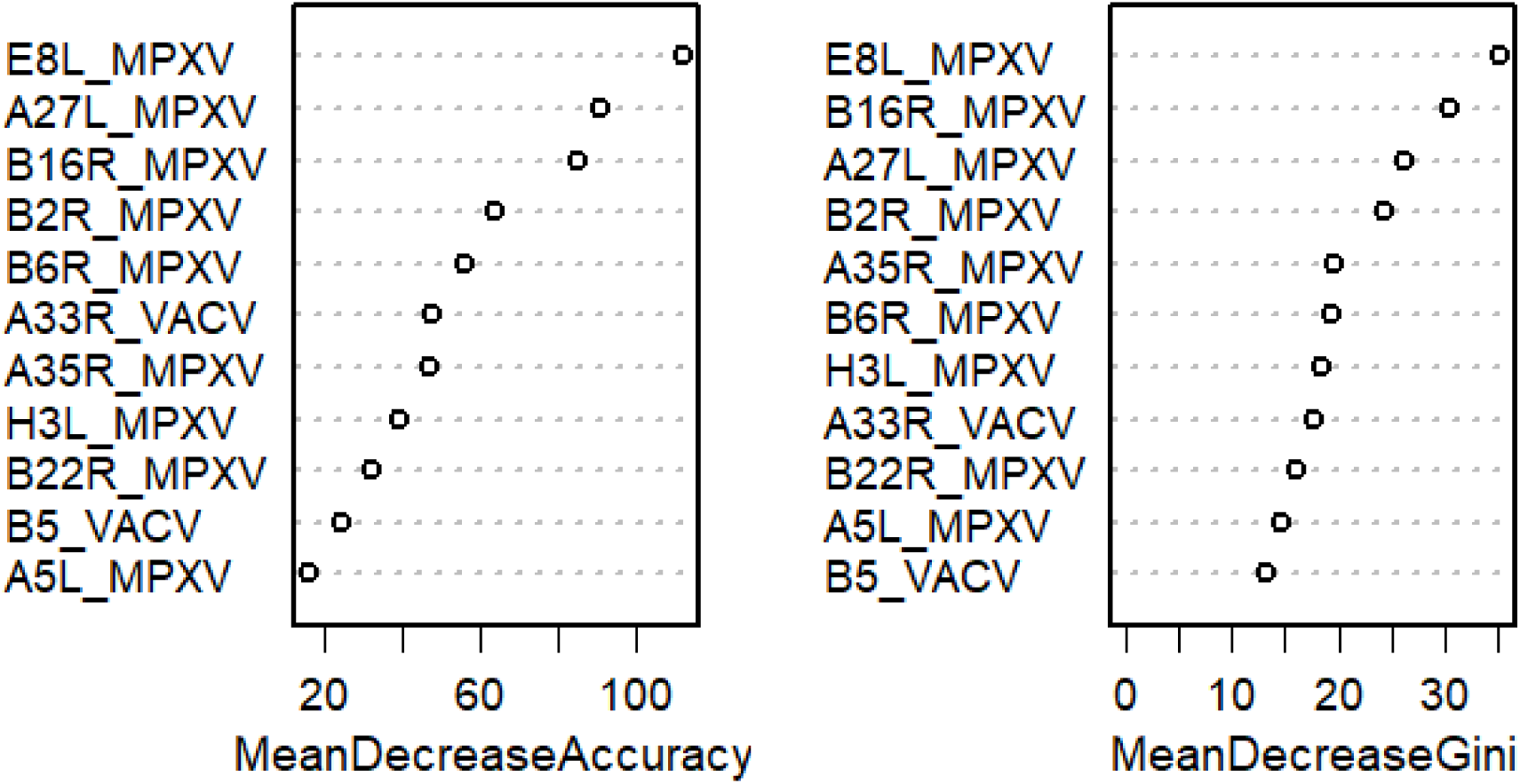
Mean decrease in accuracy (MDA) and Gini for different antigens in the IgG Luminex assay for time since infection model. Left: MDA when comparing MPXV infected vs. negative control samples from individuals born after 1976 (smallpox vaccine-naïve); right: MDA when comparing MPXV infected vs. negative control samples from individuals born in or before 1976 (smallpox vaccine-naïve)

